# An Open, Reproducible Gamma-Variate Pipeline for CT-Perfusion Time–Attenuation Curve Analysis, with Standardized (ASIST-Japan) Map Visualization

**DOI:** 10.64898/2026.06.26.26356666

**Authors:** Shuji Yamamoto

**Affiliations:** Institute of One (supported by Lisit Co., Ltd., Japan; TexelCraft OÜ, Estonia)

## Abstract

CT perfusion (CTP) is central to acute-stroke and oncologic imaging, yet quantitative outputs vary substantially across vendor software, undermining reproducibility. We present an open, transparent core (ctp-core) that fits first-pass time–attenuation curves with a gamma-variate model, derives perfusion indices (peak enhancement, time-to-peak [TTP], bolus-arrival time [BAT], area under the curve [AUC]) analytically from the fitted parameters, and renders parametric maps with the ASIST-Japan standardized lookup table (a-LUT) so that visualization is comparable across sites. Every parameter, bound, and processing step is exposed. The method is validated on Monte-Carlo synthetic curves with known ground truth; no confidential or patient data are used. Across signal-to-noise ratio (SNR) levels 5–100 (200 independent runs per level) the pipeline recovers peak time to within 0.03–0.52 s and peak amplitude to within 0.4–8.1% (mean absolute error), degrading monotonically with noise; at a representative SNR of 20 it recovers peak time within 0.13 s, peak amplitude within 2.0%, and BAT within 0.51 s, with fit quality R-squared = 0.98. The reproducibility demonstration is deterministic (fixed seed) and re-runs to bit-stable metrics. All code, the synthetic-data generator, the standardized-visualization module, evaluation scripts, and a 34-test suite are released openly for independent verification. The contribution is a fully open, parameter-transparent gamma-variate plus standardized-visualization pipeline with reproducible synthetic benchmarks — a reference others can audit, reuse, and build on.

## 1. Background and Motivation

CTP-derived maps guide reperfusion decisions and are recommended in stroke guidelines, but their quantitative reliability is limited by post-processing. Identical source data processed by different commercial packages yield significantly different perfusion maps and values, and differences in deconvolution algorithms, their implementations, and thresholds materially change estimated lesion sizes [4,5]. This opacity is a reproducibility problem: clinicians and researchers cannot inspect or audit the black box.

Gamma-variate (GV) fitting is a long-established way to model first-pass tracer dynamics [1,2], often used to suppress recirculation prior to deconvolution. Display color scales are a second, under-appreciated source of cross-site variability: the same quantitative map can look entirely different depending on the lookup table, harming inter-reader and inter-institutional comparison. Open, transparent, reproducible implementations that combine documented GV fitting with a *standardized* visualization are scarce.

### Contribution

We provide an open GV-fitting and analytic-index pipeline that is (a) parameter-transparent, (b) validated on synthetic data with ground truth, (c) standardized in visualization via the ASIST-Japan a-LUT (quantitative values never altered), and (d) fully reproducible — a reference others can audit, reuse, and build on.

## 2. Methods

### 2.1 Gamma-variate model

First-pass curves are modeled with the gamma-variate function

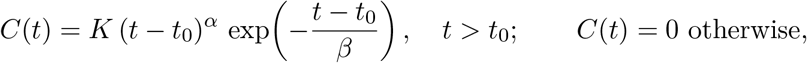

with onset *t*_0_, shape *α*, scale *β*, and scale coefficient *K*. The function is evaluated in log-space — log *C* = log *K* + *α* log(*t*− *t*_0_) − (*t* −*t*_0_)*/β* — for numerical stability, returning 0 for non-positive arguments and non-finite values. Rather than reparametrizing in terms of (*t*_max_, *y*_max_), we keep the (*K, t*_0_, *α, β*) form and obtain the peak and area analytically from the fitted parameters, which is exact and avoids a second optimization:

- Peak time: *t*_peak_ = *t*_0_ + *αβ*
- Peak value: *y*_peak_ = *K* (*αβ*)^*α*^ *e*^−*α*^
- AUC (0 → ∞): *K β*^*α*+1^ Γ(*α* + 1)
- BAT: *t*_0_ (bolus-arrival time)

### 2.2 Fitting

Baseline-corrected enhancement curves are fit by nonlinear least squares `scipy.optimize.curve_fit`, Trust Region Reflective) with physically motivated bounds: *K* ∈ [10^−6^, max(peak *·* 10^6^, 10^6^)], *t*_0_ ∈ [*t*_start_ −*T, t*_end_], *α* ∈ [0.3, 20], *β* ∈ [10^−3^, 4*T*0], where *T* is the observation window. Initialization is data-driven: *t*_0_ is placed one sample before the first crossing of 10% of the observed peak; *α* = 2 (typical bolus); *β* = (*t*_peak_− *t*_0_)*/α*; and *K* is back-solved from the peak-value identity above. The optimizer runs to `maxfev = 5000`. Fit quality is reported as R-squared and RMSE. Failures are never silent: a result object always carries a success flag, parameters, and an explicit error message (shape mismatch, too-few-samples, all-NaN, no-bolus, optimizer failure, or non-finite parameters).

### 2.3 Automatic peak detection

Two complementary index paths are provided. (i) A fit-free raw path: TTP is the time of the curve maximum; peak is the curve maximum; AUC is the trapezoidal integral of the positive part; BAT is the first time the curve exceeds 10% of its peak on the rising limb; the path returns NaN (never an exception) when the curve is unusable. (ii) The fit-derived path (Section 2.1) returns peak/TTP/AUC/BAT analytically from (*K, t*_0_, *α, β*), which is robust to per-sample noise because it uses the whole curve. For voxel-wise maps, either path can be applied across a brain mask, with a per-voxel failure mask and an overall success rate.

### 2.4 Arterial input function and derived perfusion maps

For full parametric mapping the pipeline detects the arterial input function (AIF) by a three-stage procedure — candidate screening (top-5% peak, earliest-40% TTP, baseline −50 to 200 HU to exclude bone/air), shape ranking (score = peak + wash-in-slope − FWHM −TTP, all min-max normalized), and spatial connected-component clustering — then deconvolves tissue curves against the AIF by block-circulant (delay-insensitive) truncated singular-value decomposition (threshold 15% of the largest singular value) to produce CBF, CBV (central-volume theorem, MTT = CBV/CBF *×* 60), TTP, and Tmax (AIF selection and deconvolution follow [6–11]). The DICOM workflow that feeds this stage lives in a separate private application (Section 3) and is out of scope for the open core’s reproducible claims in this preprint.

### 2.5 Standardized visualization (ASIST-Japan a-LUT)

Parametric maps are rendered with the ASIST-Japan standard a-LUT, a 256-level RGB table mapping low-to-high values through black–blue–cyan–green–yellow–red [12]. Scalars are linearly windowed to standard display ranges (CBF 0–80 mL/100 g/min, CBV 0–8 mL/100 g, MTT 0–12 s, TTP 0–25 s, Tmax 0–14 s), normalized, and indexed (idx = round(norm × 255)). Color mapping affects visualization only; quantitative voxel values are preserved unchanged and stored separately. The mapping is deterministic: identical scalar values yield bit-exact RGB, verified by unit tests; grayscale and custom research LUTs are also supported.

## 3. Implementation and scope boundary

The open core, `ctp-core` v0.1.0, is pure Python (>= 3.9) depending only on `numpy, scipy`, and `matplotlib`. It is standalone and self-contained: no graphical user interface, no DICOM input/output, and no patient or client data. Package modules: `gamma_fit, preprocessing`, `tdc_analysis, aif_detection`, `parametric_maps, synthetic, a_lut`, plus the packaged ASIST LUT asset. The interactive GUI application (DICOM workflow, viewer, batch processing) is a separate, private project and is intentionally not part of the open core: the reusable method is open, while the productized, patient-data-handling application is not.

## 4. Validation

### 4.1 Synthetic (ground-truth) experiments

Monte-Carlo curves are generated from known GV parameters (amplitude 60, *t*_0_ = 8 s, *α* = 3, *β* = 2, 40 samples at *dt* = 1 s; analytic ground truth *t*_peak_ = 14 s, *y*_peak_ = 60, BAT = 8 s) with additive Gaussian noise at controlled SNR and fixed seeds. The reference single-curve demonstration (seed 0, SNR 20) recovers *t*_peak_ = 14.14 s, peak = 57.54, BAT = 8.21 s at R-squared = 0.988 (absolute errors 0.14 s, 2.46, 0.21 s respectively); the fit-free raw path gives TTP = 13.0 s, peak = 56.59 (Figure 1).

**Figure 1.**
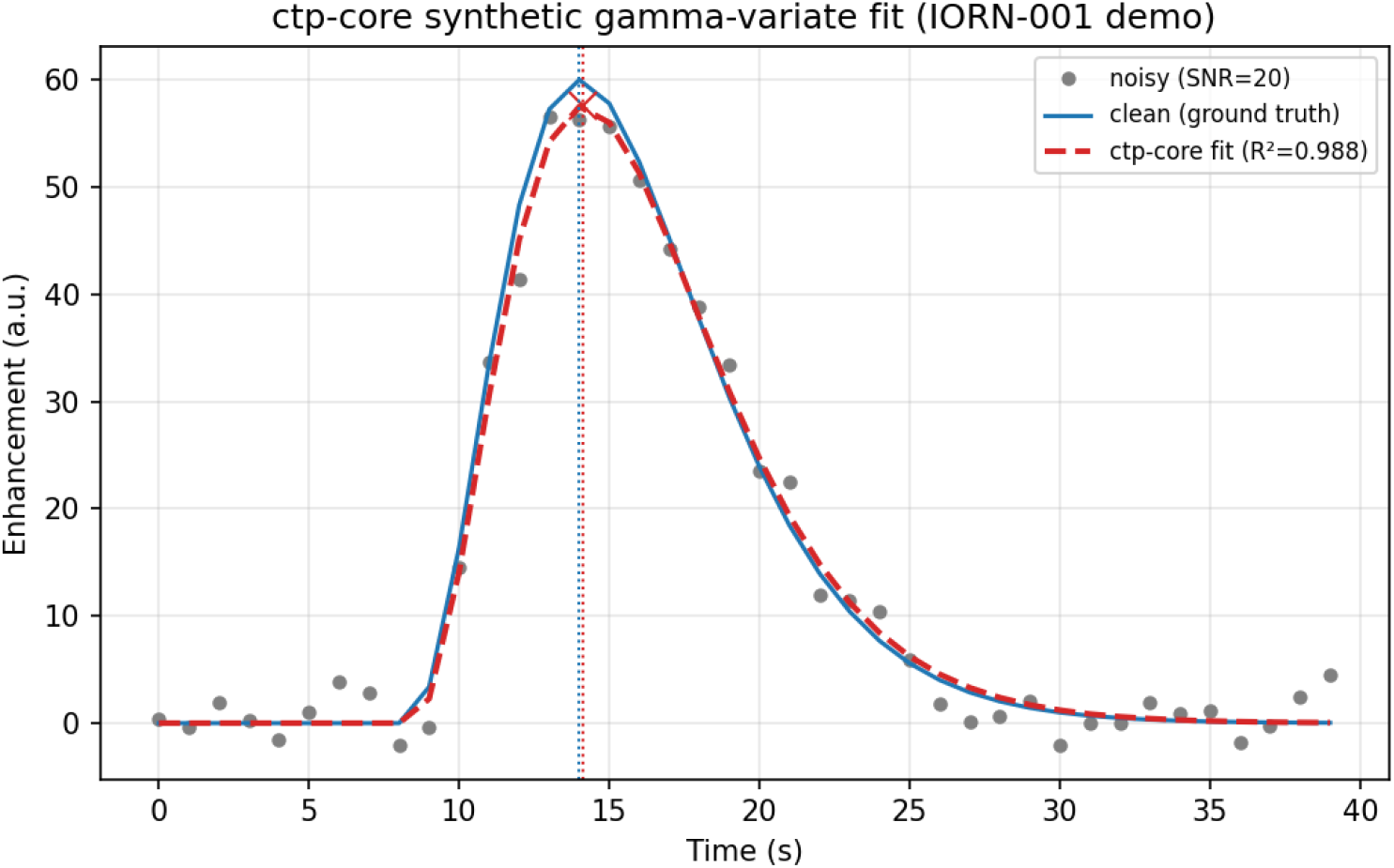
Clean (ground-truth) curve, noisy observation (SNR 20), and ctp-core gamma-variate fit with recovered peak; deterministic (seed 0).

A sweep of 200 independent noise realizations per SNR level quantifies the recovery–noise trade-off (Table 1, Figure 2). Errors degrade monotonically and predictably with noise, and the fit-derived peak is consistently closer to ground truth than the raw maximum.

At SNR 5, one of 200 runs failed the fit-quality gate and is excluded — surfaced explicitly, not hidden.

**Table 1.** Gamma-variate recovery versus SNR (mean ± SD over 200 runs per level).

| SNR | n | abs. $\Delta t_{\text{peak}}$ (s) | rel. $\Delta_{\text{peak}}$ (%) | abs. $\Delta_{\text{BAT}}$ (s) | R-squared |
| --- | --- | --- | --- | --- | --- |
| 5 | 199 | $0.518 \pm 0.381$ | $8.08 \pm 5.85$ | $2.150 \pm 1.984$ | $0.752 \pm 0.059$ |
| 10 | 200 | $0.259 \pm 0.196$ | $4.05 \pm 2.85$ | $1.137 \pm 1.143$ | $0.924 \pm 0.019$ |
| 20 | 200 | $0.128 \pm 0.102$ | $2.03 \pm 1.42$ | $0.513 \pm 0.450$ | $0.980 \pm 0.005$ |
| 40 | 200 | $0.063 \pm 0.049$ | $1.02 \pm 0.71$ | $0.245 \pm 0.199$ | $0.995 \pm 0.001$ |
| 100 | 200 | $0.025 \pm 0.020$ | $0.41 \pm 0.29$ | $0.097 \pm 0.078$ | $0.999 \pm 0.000$ |

**Figure 2.**
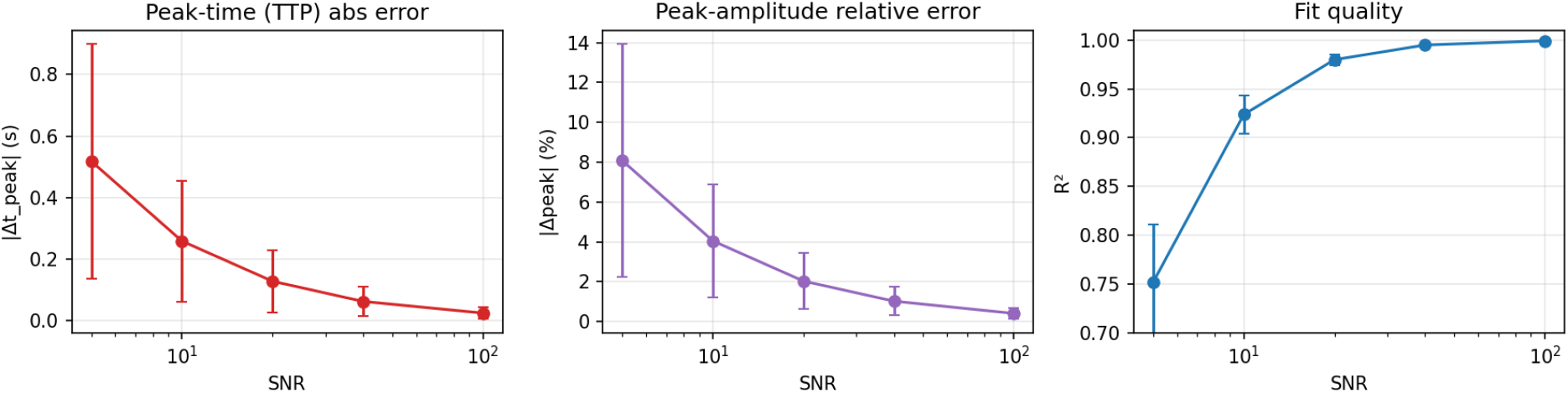
Recovery error versus SNR (200 Monte-Carlo runs per level): peak-time absolute error, peak-amplitude relative error, and fit R-squared. Error bars are *±* SD.

### 4.2 Standardized visualization demonstration

The a-LUT module is applied to a deterministic synthetic perfusion phantom (central ischemic core). Grayscale and ASIST a-LUT renderings of the identical quantitative map are shown side by side (Figure 3); the standardized palette makes the low-CBF core immediately legible while the underlying voxel values are unchanged.

**Figure 3.**
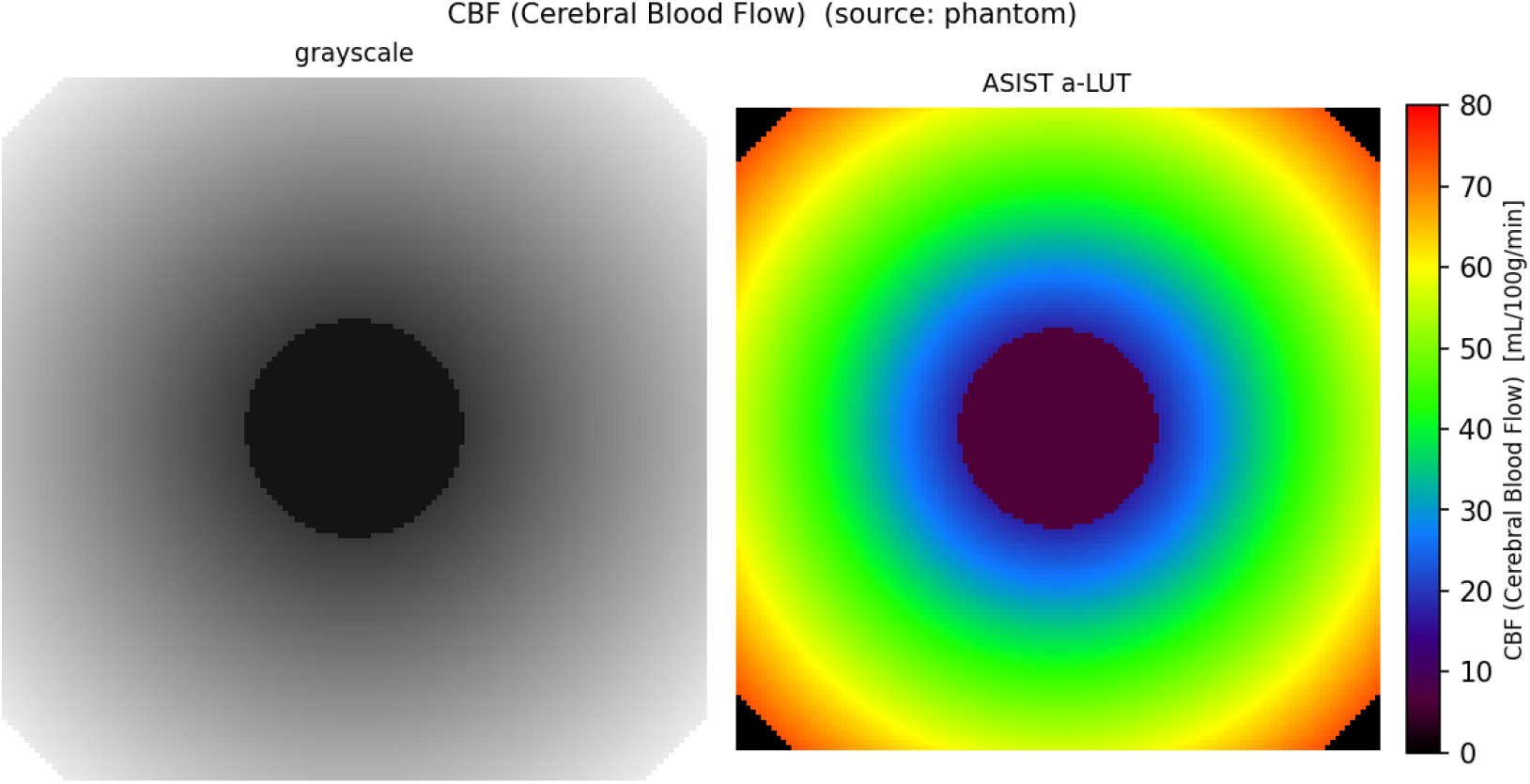
Identical synthetic CBF map rendered in grayscale versus ASIST-Japan a-LUT (standard 0–80 mL/100 g/min window). Visualization only; quantitative values preserved.

## 5. Reproducibility

All results are reproducible from the released code. Environment: Python >= 3.9 with `numpy >= 1.21, scipy >= 1.7, matplotlib >= 3.5.` The single-curve demonstration `(examples/run_synthetic_demo.py`) and the SNR sweep producing Table 1 and Figure 2 `(examples/run_snr_sweep.py`, seeds 0–199 per level) are deterministic; the a-LUT figures are produced by `make_a_lut_figures.py;` the test suite `(pytest tests/`) reports 34 passing tests. At seed 0, SNR 20, the peak-time absolute error is 0.140 s and R-squared is 0.988; Table 1 reproduces to the reported mean ± SD.

## 6. AI-Use Disclosure

This manuscript and the associated software were produced by a human author (S. Yamamoto), who is solely accountable for their content. AI agents were used as tools: code scaffolding and refactoring of the scientific modules, test drafting, figure and script generation, and manuscript drafting were assisted by a large language model (Claude, Anthropic). The author independently re-executed every numerical result reported here (the demonstration, the SNR sweep, and the test suite) and verified all figures, equations, and claims against the code. No AI system is an author. This disclosure follows ICMJE and COPE guidance: AI is reported as a tool, not credited with authorship.

## 7. Limitations

Gamma-variate is a first-pass model, not full deconvolution; the open-core reproducible claims here are established on synthetic data with single-AIF assumptions, across the stated SNR range and one curve geometry. The full CBF/CBV/MTT mapping and AIF detection (Section 2.4) are documented and implemented but are exercised through the private DICOM application and are not part of this preprint’s reproducible benchmark. No clinical, regulatory, or external real-data validation is claimed; external validation on a public dataset (for example ISLES’18/’24) and broader parameter sweeps are planned. As released, `ctp-core` is a research reference, not a clinical or regulatory-grade tool.

## Data Availability

All data in this study are synthetic and are reproducibly generated by the openly released code. All code, the synthetic-data generator, the standardized-visualization module, and evaluation scripts are available online at https://github.com/Institute-of-One/ctp-core under the MIT license, archived on Zenodo (DOI 10.5281/zenodo.20921268).

https://doi.org/10.5281/zenodo.20921268

## Declarations

### Data and code availability

All code, the synthetic-data generator, the standardized-visualization module, evaluation scripts, and the test suite are openly available at https://github.com/Institute-of-One/ctp-core under the MIT license, archived on Zenodo (all-versions DOI 10.5281/zenodo.20921268 v0.1.0 DOI 10.5281/zenodo.20921269). No patient, clinical, or client data were used; all data in this study are synthetic and produced by the included reproducible generator.

### Ethics

Not applicable. This study involved no human participants, animal subjects, or patient data; only synthetic data were analyzed.

### Competing interests

The author conducts commercial imaging-analysis services through Lisit Co., Ltd. (Japan) and TexelCraft OÜ (Estonia), which support the Institute of One research brand. This work used no client or patient data and presents an openly licensed method. The author declares no other competing interests.

### Funding

This work received no specific grant from any funding agency in the public, commercial, or not-for-profit sectors.

### Author contributions

S.Y. is the sole author and is responsible for conceptualization, methodology, software, validation, formal analysis, visualization, and writing. AI tools were used as disclosed in Section 6.

## Notes

### Competing Interest Statement

The author conducts commercial imaging-analysis services through Lisit Co., Ltd. (Japan) and TexelCraft OU (Estonia), which support the Institute of One research brand. This work used no client or patient data and presents an openly licensed method. The author declares no other competing interests.

